# The emergence of COVID-19 in Indonesia: analysis of predictors of infection and mortality using independent and clustered data approaches

**DOI:** 10.1101/2020.07.10.20147942

**Authors:** Erlina Burhan, Ari Fahrial Syam, Ahmad Jabir Rahyussalim, Prasenohadi, Navy G Lolong Wulung, Agus Dwi Susanto, I Gede Ketut Sajinadiyasa, Dewi Puspitorini, Dewi Lestari, Indah Suci Widyahening, Vivi Setiawaty, Dwiana Ocviyanti, Kartika Qonita Putri, Aswin Guntara, Davrina Rianda, Anuraj H Shankar, Rina Agustina

## Abstract

**Background:** Analyses of correlates of SARS-CoV-2 infection or mortality have usually assessed individual predictors. This study aimed to determine if patterns of combined predictors may better identify risk of infection and mortality

**Methods:** For the period of March 2^nd^ to 10^th^ 2020, the first 9 days of the COVID-19 pandemic in Indonesia, we selected all 18 confirmed cases, of which 6 died, and all 60 suspected cases, of which 1 died; and 28 putatively negative patients with pneumonia and no travel history. We recorded data for travel, contact history, symptoms, haematology, comorbidities, and chest x-ray. Hierarchical cluster analyses (HCA) and principal component analyses (PCA) identified cluster and covariance patterns for symptoms or haematology which were analysed with other predictors of infection or mortality using logistic regression.

**Results:** For univariate analyses, no significant association with infection was seen for fever, cough, dyspnoea, headache, runny nose, sore throat, gastrointestinal complaints (GIC), or haematology. A PCA symptom component for fever, cough, and GIC tended to increase risk of infection (OR 3.41; 95% CI 1.06-14; p=0.06), and a haematology component with elevated monocytes decreased risk (OR 0.26; 0.07-0.79; 0.027). Multivariate analysis revealed that an HCA cluster of 3-5 symptoms, typically fever, cough, headache, runny nose, sore throat but little dyspnoea and no GIC tended to reduce risk (aOR 0.048; <0.001–0.52; 0.056). In univariate analyses for death, an HCA cluster of cough, fever and dyspnoea had increased risk (OR 5.75; 1.06 − 31.3, 0.043), but no other individual predictor, cluster or component was associated. Other significant predictors of infection were age ≥ 45, international travel, contact with COVID-19 patient, and pneumonia. Diabetes and history of contact were associated with higher mortality.

**Conclusions:** Cluster groups and co-variance patterns may be stronger correlates of SARS-CoV-2 infection than individual predictors. Comorbidities may warrant careful attention as would COVID-19 exposure levels.

## Introduction

The coronavirus SARS-CoV-2 is a deadly respiratory pathogen first reported in Wuhan City, Hubei Province of China in early 2020, and rapidly spread globally. On 11 March 2020, the World Health Organization declared a pandemic(1), and by 2 May 2020 the number of confirmed cases worldwide reached nearly 3.3 millions with more than 230,000 deaths, and a global case fatality rate of 7.04%.(2) Indonesia, a large archipelago country in South-east Asia with a population of 260 million, did not report a confirmed COVID-19 case until 2 March 2020, several weeks after a total of 217 cases had been reported from neighboring countries of Singapore, Malaysia, Thailand and Australia. This raised concerns from groups both inside and outside the country regarding diagnostic and reporting practice. However, by 2 May 2020 the number of confirmed cases were 10,551, with 800 deaths, yielding a case fatality rate of 7.58%, amongst the highest in the world.(2) Again, this raised concerns about both the diagnostic capacity and care of cases.

The availability of routine data for case history, symptoms, haematology and radiology presents an opportunity to define presentation patterns predictive for COVID-19. Knowledge of anomalous risk patterns may be especially useful at the early phase of the epidemic in new locations when high throughput molecular testing is not yet available. Analyses of syndromic and haematology data in some studies showed the most common symptoms of COVID-19 were fever, cough, and lymphocytopenia. Patients may also be asymptomatic yet with abnormal findings from lung imaging.(3) Previous analyses typically examined each symptom, exposure or hematological characteristic individually or as separate predictors of SARS-CoV-2 infection, pathogenicity, or mortality. Few studies have examined clusters or co-variance patterns of symptoms, exposures, haematological data, or combinations thereof, and their associations with outcomes.

We therefore analysed routine clinical data and patient history from all hospital admissions during the first nine days of the epidemic in Indonesia who were confirmed by real time polymerase chain reaction (RT-PCR) as COVID-19 cases, and all suspect but virus negative cases, and other lung infection cases presumed negative for the virus. We applied hierarchical cluster analysis (HCA) and principal components analysis (PCA) to discern specific clusters or co-variance patterns of syndromic and haemotological data, and analysed these along with radiological features and patient history for associations with SARS-CoV-2 infection or death using univariate and multivariate logistic regression. These analytic approaches may prove useful for more rapid recognition of emergent COVID-19 cases, thereby hastening treatment, and enhancing survival.

## Materials and methods

### Study setting

This was a retrospective cohort study of patients from four collaborating hospitals in Jakarta, Bali and Jambi provinces which had reported suspected and confirmed case of COVID-19 to the Ministry of Health. We collected data from confirmed, suspected and lung infection cases admitted from March 2^nd^ to 10^th^ 2020, the first nine days of cases in Indonesia. Ethical approval was obtained from the Research Ethics Committee of the Faculty of Medicine, Universitas Indonesia – Dr. Cipto Mangunkusumo General Hospital, with approval number 20030331. After consideration of ethical issues, logistics and urgency of this work, the Committee waived the requirement for written individual informed consent and approved the sharing of anonymized data.

### Patients identification

As per hospital protocols, patients admitted with respiratory infections had been registered as potential COVID-19 cases and been classified as under-supervision or monitored. Under-supervision had included patients with clinical symptoms of fever (>38 °C) or history of fever, runny nose, sore throat, and pneumonia-like symptoms such as cough, dyspnoea, sweating, sharp chest pain worsened by breathing deeply, or cough with a history of travel to a COVID-19 affected country in the last 14 days, or having contact with a person with acute upper respiratory tract infection. Monitored had been defined as upper respiratory symptoms without pneumonia-like symptoms, plus travel history to an affected country within 14 days.

A confirmed COVID-19 case was an under-supervision or monitored patient with a positive SARS-CoV-2 test by RT-PCR for naso-oropharyngeal swab, sputum, or bronchoalveolar lavage (18 patients); a suspected case was a patient twice-confirmed negative by RT-PCR test (47 patients) or not assessed (13 patients); and formal classification required the case had been reported to the Ministry of Health of the Republic Indonesia. For comparison, we selected patients with community acquired pneumonia who were admitted between January 1^st^ to March 10^th^ 2020 and reported no history of international travel. A total of 18 confirmed COVID-19 patients, 60 suspected patients, and 28 lung patients were identified.

### Data collection

Study physicians reviewed medical records of hospitalized patients, and in accord with World Health Organization diagnostic guidelines, and hospital procedures, data were extracted for demographics, nationality, travel history, clinical signs, radiological features, and haemotology, including haemoglobin and hematocrit, and counts for thrombocytes, total leucocytes, monocytes, lymphocytes, basophils, neutrophils, and eosinophils. Patients with symptoms of acute respiratory tract infection or travel history from COVID-19-affected countries within the last 14 days had been assessed in the emergency room or out-patient clinic(4), and case history, physical examination and chest x-rays had been done within the hospital. Venous blood samples had been taken for complete white blood cell count. Some samples were assessed for electrolytes, lactic acid products, C-reactive protein, procalcitonin, and liver as well as renal function tests; these tests were selectively assessed and gaps in data precluded inclusion in this analysis. Chest computed tomography (CT) scan was not reported for any patient.(5)

Naso-oropharyngeal swab, sputum or bronchoalveolar lavage specimens had been sent to the National Institutes for Health Research and Development (NIHRD) laboratory in Jakarta for RT-PCR to assess presence of the virus. Tests were performed after virus inactivation, and RNA was extracted using the QiAmp RNA viral kit based on the instructions and standard laboratory techniques, followed by RT-PCR using SuperScript™ III Platinum® One-Step Quantitative RT-PCR for the N gene in accord with US CDC protocols.(6)

### Statistical analyses

Descriptive statistics for continuous data are presented as the mean and standard deviation (SD) for normally distributed data, and median and interquartile range (IQR) for non-normal data. Normality of haematology data was assessed by the Shapiro-Wilk test and QQ plots and, if needed, data were normalized by log, square root, or cube root transforms, or reflection followed by transform. Normalized data were standardized to a mean of zero and standard deviation of one. Comparisons for differences between groups was by one-way ANOVA or the Kruskal-Wallis test for non-parametric data. Categorical and binary data were expressed as the number and proportion (%) with comparison using the χ^2^ test. Univariate logistic regression was used to identify the association of risk factors, or independent variables, such as demography, symptoms, comorbidities, chest x-ray abnormalities, and laboratory assessments, with SARS-CoV-2 infection or death as dependent variables.

Multivariate logistic regression with backward selection was used with a cutoff of p<0.15 from univariate regression, and retained variables previously reported to affect the outcomes. Firth’s logistic regression was used to permit calculation of regression coefficients when standard logistic regression failed due to the binary outcome prevalence being very high or very low for a particular risk factor. A two-sided α of <0.05 was considered statistically significant. SPSS V.26.0 and SAS 9.4 were used for all analyses.

PCA was performed to identify composite symptom and laboratory components based on their co-variance constructs. Categorical PCA was used for binary symptom data, and conventional PCA for continuous, normalized, and standardized laboratory data. For laboratory data, missing values were imputed as the mean of each variable. A component was retained following cross validation if eigenvalues were above the cutoffs defined by Horn’s parallel analysis. Principal component scores were recoded to tertiles and used as predictors of SARS-CoV-2 infection or death by logistic regression with the lowest tertile as the reference category.

HCA was used to define clusters of association of symptoms and other characteristics. The number of retained clusters were determined as 5 based on the highest adjusted Rand index with a high number of clusters and a high pseudo T2 statistic. Cluster memberships were used as predictors of SARS-CoV-2 infection or death by logistic regression.

Symptoms and hematological data were analysed as separate variables as risk factors for associations with SARS-CoV-2 infection or death using univariate and multivariate logistic regression, and after classification into patterns or clusters of risk factors defined by PCA or HCA, followed by logistic regression.

## Results

In Table 1 we summarize patient characteristics, including age, gender, history of international exposure, history of contact, symptoms, haematology, comorbidities, chest x-ray abnormalities, and diagnosis at discharge. A total of 106 subjects comprised of 18 confirmed COVID-19, 60 suspected, and 28 lung infection cases. The mean age of all subjects was 44.8 ± 18.7 years with 42% being 45-65 years old. The mean age of confirmed cases was 52.6 ±10.4 years, older than suspected patients who were 35.6 ± 17.3 years, but younger than for lung infection cases who were 59.3 ± 14. The gender distribution was nearly equal, except in the confirmed case group wherein 66.7% were male. International travel history was found in 50% of confirmed, and 85.0% of suspected cases. Moreover, 50% of confirmed and 16.7% of suspected cases had contact history with COVID-19 patients.

**Table 1.** General characteristics of the study patients by category.

| Characteristics | All | Non COVID-19 |  | COVID-19 | Survivor | Death |
| --- | --- | --- | --- | --- | --- | --- |
|  |  | Suspected | Lung infection | Confirmed |  |  |
|  | n= 106 | n = 60 | n=28 | n = 18 | n=99 | n=7 |
| **Demography** |  |  |  |  |  |  |
| Age (yr)<sup>a</sup> | 44.8 (18.7) | 35.6 (17.3) | 59.3 (14.0) | 52.6 (10.4) | 43.5 (18.6) | 59.6 (6.29) |
| Distribution (yr), n (%) |  |  |  |  |  |  |
| 0-18 | 8 (7.5) | 8 (13.3) | 0 (0) | 0 (0) | 8 (8.0) | 0 (0) |
| 19-30 | 13 (12.3) | 11 (18.3) | 1 (3.6) | 1 (5.6) | 13 (13.0) | 0 (0) |
| 31-44 | 29 (27.4) | 23 (38.3) | 3 (10.7) | 3 (16.7) | 29 (29.0) | 0 (0) |
| 45-65 | 45 (42.5) | 16 (26.7) | 16 (57.1) | 13 (72.2) | 39 (39.0) | 7 (100) |
| > 65 | 11 (10.4) | 2 (3.3) | 8 (28.6) | 1 (5.6) | 11 (11.0) | 0 (0) |
| Male, n (%) | 55 (51.9) | 29 (48.3) | 14 (50.0) | 12 (66.7) | 51 (51.0) | 4 (57.1) |
| International travel, n (%) | 59 (55.7) | 51 (85.0) | 0 (0) | 8 (44.4) | 59 (59.6) | 1 (14.3) |
| No | 47 (44.3) | 9 (15.0) | 28 (100) | 10 (55.6) | 42 (42.0) | 6 (85.7) |
| China | 17 (16.0) | 17 (28.3) | 0 (0) | 0 (0) | 17 (17.0) | 0 (0) |
| Singapore and/ Malaysia | 19 (17.9) | 14 (23.3) | 0 (0) | 5 (27.8) | 18 (18.0) | 1 (14.3) |
| Asian countries | 17 (16.0) | 16 (26.7) | 0 (0) | 1 (5.6) | 17 (17.0) | 0 (0) |
| Other countries | 5 (4.7) | 4 (6.7) | 0 (0) | 1 (5.6) | 5 (5.0) | 0 (0) |
| Diamond Princess Boat | 1 (0.9) | 0 (0) | 0 (0) | 1 (5.6) | 1 (1.0) | 0 (0) |
| Contact history, n (%) | 19 (17.9) | 10 (16.7) | 0 (0) | 9 (50.0) | 14 (14.1) | 5 (71.4) |
| **Symptoms<sup>b</sup>, n (%)** |  |  |  |  |  |  |
| Fever duration (days)<sup>a</sup> | 3 (1 - 5.3) | 2 (1 - 3) | 3 (3 - 6.5) | 10 (7 - 14) | 3 (1-5) | 3 (6 - 11.5) |
| Fever | 83 (78.3) | 48 (80.0) | 20 (71.4) | 15 (83.3) | 78 (78.0) | 5 (71.4) |
| Headache | 8 (7.5) | 8 (13.3) | 0 (0) | 0 (0) | 8 (8.1) | 0 (0) |
| Runny nose | 20 (18.9) | 20 (33.3) | 0 (0) | 0 (0) | 20 (20) | 0 (0) |
| Cough | 89 (84.0) | 48 (80.0) | 25 (89.3) | 16 (88.9) | 83 (83.8) | 6 (85.7) |
| Sore throat | 11 (10.4) | 11 (18.3) | 0 (0) | 0(0) | 11 (11.0) | 0 (0) |
| Dyspnoea | 51 (48.1) | 18 (30.0) | 24 (85.7) | 9 (50.0) | 45 (45.5) | 6 (85.7) |
| GIT complaints | 5 (4.7) | 5 (8.3) | 0 (0) | 0 (0) | 5 (5.1) | 0 (0) |
|  |  | Suspected | Lung infection | Confirmed |  |  |
|  | n= 106 | n = 60 | n=28 | n = 18 | n=99 | n=7 |
| **Coexisting disorder, n (%)** |  |  |  |  |  |  |
| Any | 45 (42.5) | 10 (16.7) | 27 (96.4) | 8 (44.4) | 41 (41.4) | 4 (57.1) |
| Diabetes | 14 (13.2) | 2 (3.3) | 7 (25) | 5 (27.8) | 10 (10.0) | 4 (57.1) |
| Hypertension | 13 (12.3) | 3 (5) | 5 (17.9) | 5 (27.8) | 11 (11.0) | 2 (28.6) |
| Heart disease<sup>c</sup> | 8 (7.5) | 1 (1.7) | 7 (25) | 0 (0) | 8 (8.1) | 0 (0) |
| Malignancy | 6 (5.7) | 2 (3.3) | 1 (3.6) | 3 (16.7) | 5 (5.0) | 1 (14.3) |
| Kidney Diseases | 4 (3.8) | 0 (0) | 4 (14.3) | 0 (0) | 9 (9) | 0 (0) |
| Respiratory Disorder | 19 (17.9) | 2 (3.3) | 17 (60.7) | 0 (0) | 19 (19.0) | 0 (0) |
| Combined severe diseases<sup>d</sup> | 34 (32.1) | 7 (11.7) | 24 (85.7) | 3 (16.7) | 33 (33.3) | 1 (14.3) |
| **Laboratory results** | **n =99** | **n = 53** | **n = 28** | **n = 18** | **n =93** | **n =6** |
| Haemoglobin (g/dL)<sup>a</sup> | 12.9 (2.1) | 13.1 (1.7) | 11.7 (2.4) | 13.8 (2.0) | 37.5 (5.99) | 37.6 (6.27) |
| Anaemia, n (%) | 30 (30.3) | 10 (18.9) | 4 (22.2) | 16 (57.1) | 12.8 (2.07) | 13.1 (2.23) |
| Haematocrit (%)<sup>a</sup> | 37.5 (6.0) | 38.7 (4.7) | 34 (7.0) | 39.4 (5.6) | 37.5 (5.99) | 37.6 (6.28) |
| Leucocyte (10<sup>3</sup>/μL)<sup>a</sup> | 10.3 (7.1-16.4) | 8 (5.71-11.8) | 16.1 (11.2-21.5) | 7.06 (3.88-10.3) | 10.2 (6.53-14.3) | 12.8 (8.48-15.3) |
| ≤10, n (%) | 46 (46.9) | 33 (63.5) | 4 (14.3) | 9 (50.0) | 44 (47.8) | 2 (33.3) |
| >10, n (%) | 52 (53.1) | 19 (36.5) | 24 (85.7) | 9 (50.0) | 48 (52.2) | 4 (66.7) |
| Thrombocyte (10<sup>3</sup>/μL)<sup>a</sup> | 268 (198-347) | 236 (188-320) | 320 (239-394) | 293 (210-328) | 268 (207-345) | 202 (157-215) |
| <150, n (%) | 6 (6.2) | 4 (7.8) | 1 (3.6) | 1 (5.6) | 5 (5.5) | 1(16.7) |
| 150-450, n (%) | 84 (86.6) | 45 (88.2) | 22 (78.6) | 17 (94.4) | 79 (86.8) | 5 (83.3) |
| >450, n (%) | 7 (7.2) | 2 (3.9) | 5 (17.9) | 0 (0) | 7 (7.7) | 0 (0) |
|  | **n =71** | **n = 52** | **n = 28** | **n = 18** | **n = 86** | **n = 6** |
| Lymphocyte (%) | 11.9 (8.36-24.7) | 22.6 (11.9-36.6) | 9.45 (3.83- 11.8) | 21.8 (10.4-26.4) | 14.1 (8.39-25.4) | 7 (4.70-8.75) |
| <20 | 58 (63.0) | 19 (41.3) | 26 (82.9) | 13 (72.2) | 53 (60.9) | 6 (100) |
| 20-40 | 25 (27.2) | 18 (39.1) | 5 (27.8) | 2 (7.1) | 25 (28.7) | 0 (0) |
| >40 | 9 (9.9) | 9 (19.6) | 0 (0) | 0 (0) | 9 (10.3) | 0 (0) |
| Basophil (%) | 0.4 (0.2-0.68) | 0.6 (0.31-0.99) | 0.2 (0.1- 0.4) | 0.2 (0.1-1.8) | 0.3 (0.2 - 0.6) | 0.2 (0.1- 0.28) |
| Eosinophil (%) | 0.4 (0.04-1.8) | 0.46 (0.16-1.98) | 0.15 (0-1.43) | 0 (0 - 0.8) | 0.3 (0.0-1.8) | 0.1 (0.0 - 0.25) |
| Neutrophil (%) | 76.7 (65.3-86.3) | 67.7 (52.3-78.9) | 82.7 (75.9-90.5) | 80.3 (70.8-90.0) | 76.7 (66.3-84.8) | 88.7 (86.2-91.4) |
|  |  | Suspected | Lung infection | Confirmed |  |  |
|  | n= 106 | n = 60 | n=28 | n = 18 | n=99 | n=7 |
| Monocyte (%) | 7.25 (5.4-9.95) | 7.8 (6.15-10.0) | 6.85 (4.95-11.0) | 6.4 (2.8-9.6) | 7.2 (5.4-9.5) | 3.9 (2.8-5.0) |
| **Chest radiograph abnormality, n (%)** | **n=106** | **N=60** | **N=28** | **N=18** | **N=97** | **N=7** |
| Pneumonia | 76 (71.7) | 32 (53.3) | 27 (96.4) | 17 (94.4) | 0 (0) | 7 (100) |
| Cardiomegaly | 10 (9.4) | 2 (3.3) | 7 (25.0) | 1 (5.6) | 9 (9.9) | 0 (0) |
| Within normal | 17 (16.0) | 15 (25.0) | 1 (3.6) | 1 (5.6) | 17 (17.2) | 0 (0) |
| Consolidation | 20 (18.9) | 11 (18.3) | 8 (28.6) | 1 (5.6) | 19 (19.2) | 1 (14.3) |
| Patchy/Infiltration/ Interstitial | 76 (71.7) | 32 (53.2) | 27 (96.4) | 17 (94.4) | 69 (69.7) | 7 (100) |
| **Abnormality, n (%)** |  |  |  |  |  |  |
| Glasgow Coma Scale <15 | 0 (0) | 0 (0) | 0 (0) | 0 (0) | 0 (0) | 0 (0) |
| Respiratory Distress | 51 (48.1) | 18 (30.0) | 24 (85.7) | 9 (50.0) | 45 (45.5) | 6 (85.7) |
| Systolic Pressure <100 mmHg | 0 (0) | 0 (0) | 0 (0) | 0 (0) | 0 (0) | 0 (0) |
| **Discharge diagnosis, n (%)** |  |  |  |  |  |  |
| COVID-19 infection | 18 (17.2) | 0 (0) | 18 (100) | 0 (0) | 12 (12.1) | 6 (85.7) |
| Community Acquired Pneumonia | 47 (44.3) | 19 (31.7) | 0 (0) | 28 (100) | 47 (47.5) | 0 (0) |
| Viral Infection | 11 (18.3) | 11 (18.3) | 0 (0) | 0 (0) | 11 (11.1) | 0 (0) |
| Acute Bronchitis | 6 (5.6) | 6 (10.0) | 0 (0) | 9 (0) | 6 (6.1) | 0 (0) |
| Upper Respiratory Infection | 10 (9.4) | 10 (16.7) | 0 (0) | 0 (0) | 10 (10.1) | 0 (0) |
| Other diseases | 6 (5.4) | 6 (10.2) | 0 (0) | 0 (0) | 5 (5.0) | 1(14.3) |
| Not yet discharged | 8 (7.5) | 8 (13.3) | 0 (0) | 0 (0) | 8 (8.1) | 0 (0) |

*GIT* gastrointestinal; *TB* lung tuberculosis, including TB and other respiratory diseases.
<sup>a</sup> Presented as mean (SD); or as median (25th-75th percentile)
<sup>b</sup> Assessed at admission
<sup>c</sup> Heart disease included coronary artery disease, atrial fibrillation, congestive heart failure, and atherosclerotic heart disease.
<sup>d</sup> Severe diseases included Heart disease, Kidney diseases, Lung TB, other respiratory and severe Diseases

Fever and cough, followed by dyspnoea, were the most frequent symptoms followed by runny nose, sore throat, headache, and gastrointestinal complaints (GIC). Comorbidities were present in nearly half of patients, with combined severe diseases as the most common, followed by pre-existing respiratory disorders and diabetes mellitus. However, there were no pre-existing respiratory disorders in confirmed COVID-19 cases. The two most common disorders of confirmed cases were diabetes (27.8%) and hypertension (27.8%).

Chest x-ray showed pneumonia in 94.4% of confirmed and 53.3% of suspected cases. Among the suspected cases, 25% were within normal limits. Consolidation was less frequently reported in confirmed cases. Haematology did not show lymphopenia or thrombocytopenia in confirmed or suspected cases. However, a lower level of leucocytes was seen in suspected and confirmed cases, whereas leucocytosis was observed more frequently in 85% of the lung infection cases.

In univariate logistic regression, age ≥ 45 years, positive history of contact, and pneumonia by chest x-ray, were significantly associated with higher risk of SARS-CoV-2 infection. Among all symptoms, runny nose tended toward lower risk for infection. Hypertension and malignancy were comorbidities significantly associated with confirmed cases (Table 2). Multivariate logistic regression found that age ≥ 45 years (aOR 7.81; 95% CI 1.61-38.0), travel history to an affected area (aOR 63.2; 95% CI 4.65-858.9), history of contact with a COVID-19 positive patient (aOR 232; 95% CI 15.1-3569) and pneumonia by chest x-ray (aOR 31.5; 95% CI 1.74-571.3) were independently associated with SARS-CoV-2 infection.

**Table 2.**
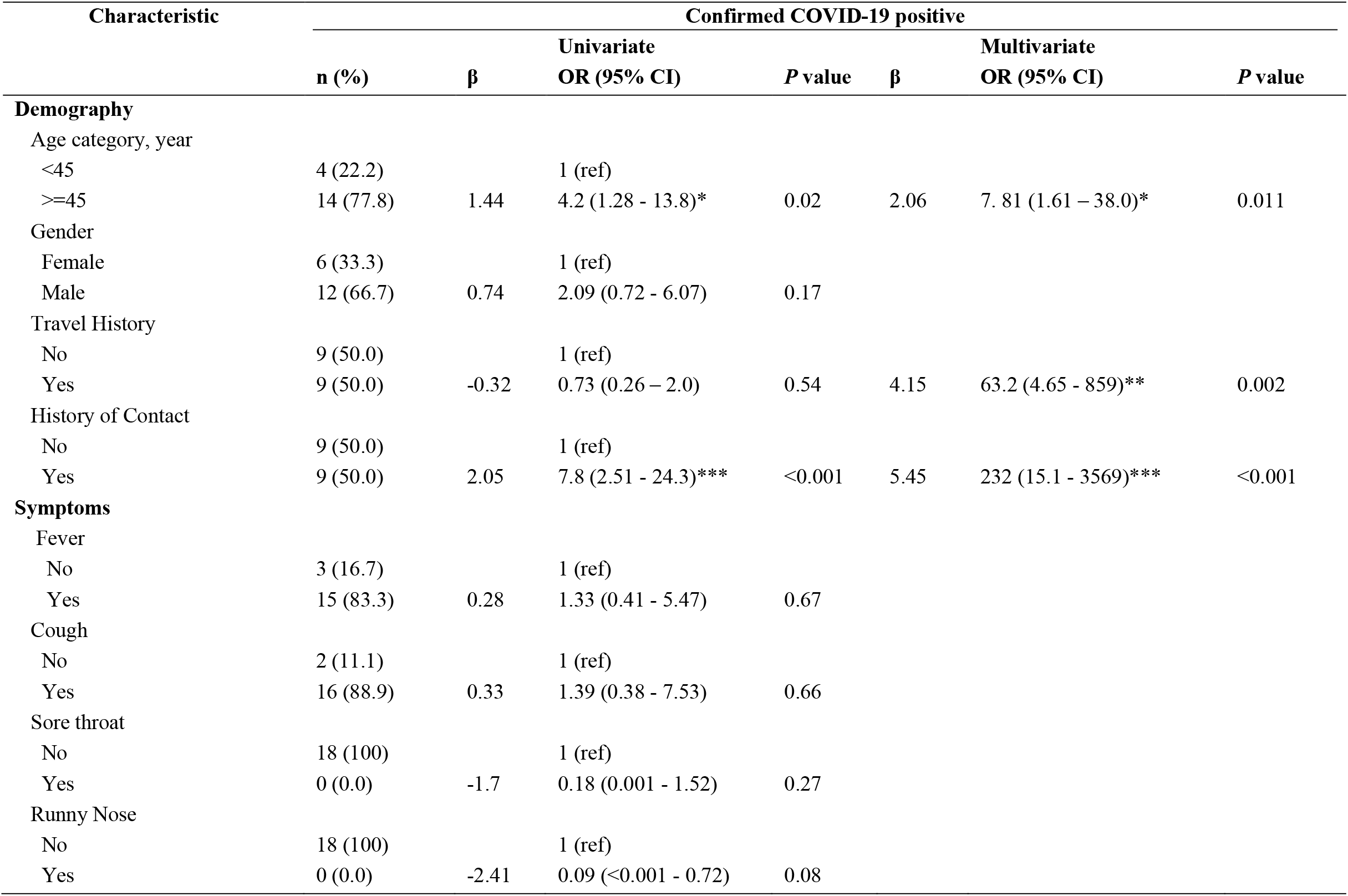
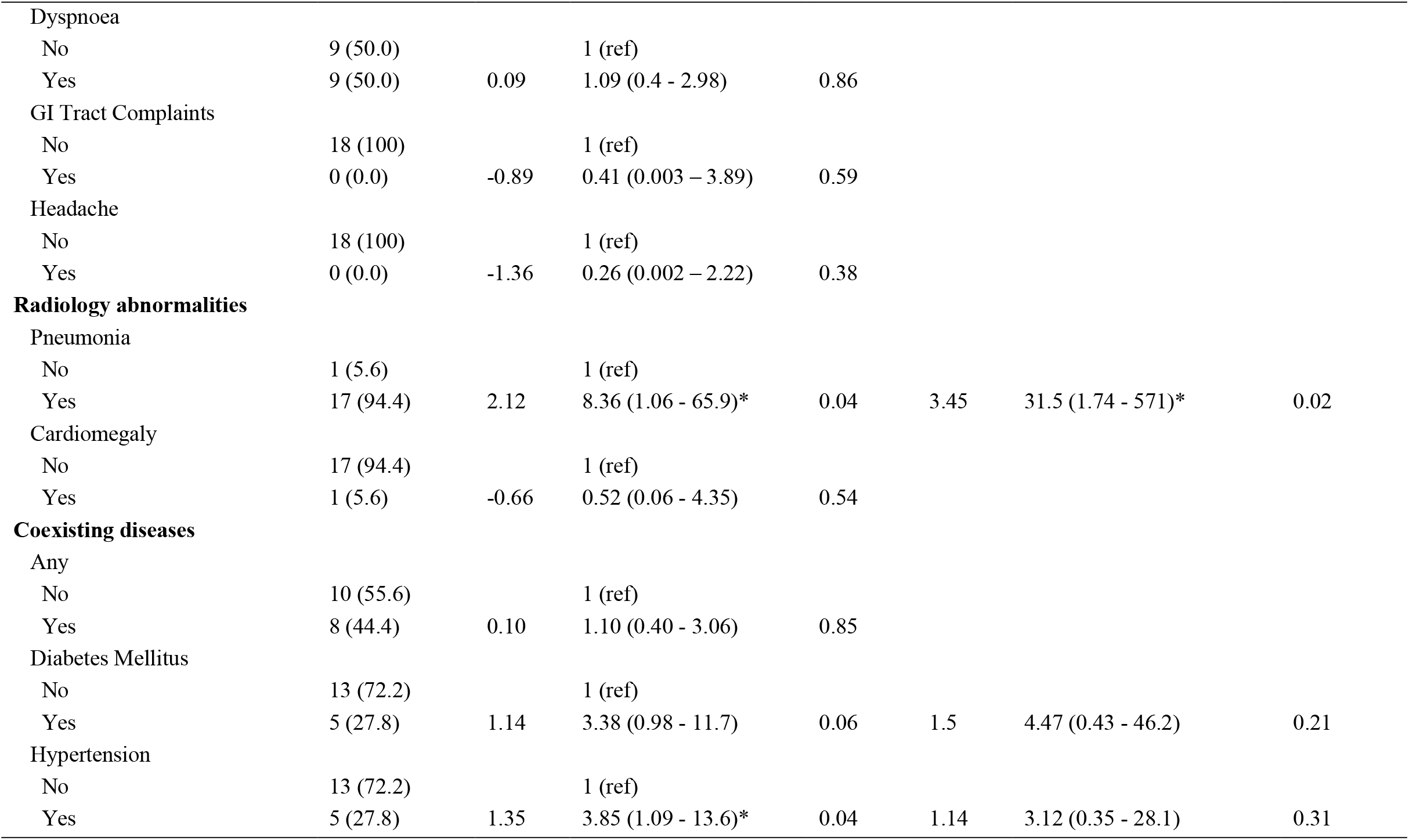
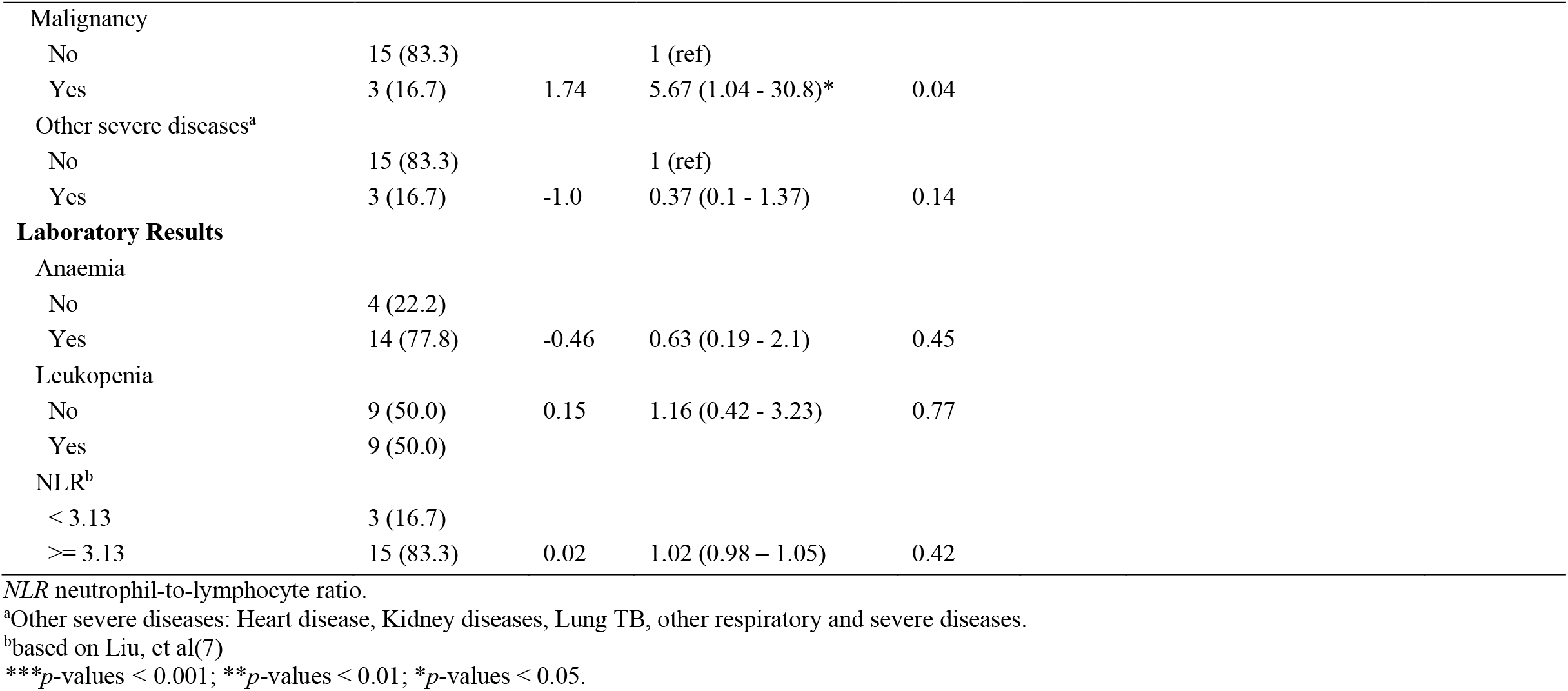
Univariate and multivariate logistic regression analyses for predictors of SARS-CoV-2 infection (n=106)

Among all patients, including 7 deaths and 99 survivors, univariate analysis (Table 3) showed that age ≥ 45 years, international travel, contact with confirmed COVID-19 case, and diabetes mellitus were associated with death. Dyspnoea tended toward increased risk for death. In the multivariate logistic regression, travel history, presence of history of contact with confirmed COVID-19 cases, and diabetes mellitus retained significant association with mortality. No significant associations were observed between laboratory markers and death. Gender was not associated with SARS-CoV-2 infection or death.

**Table 3.** Univariate and multivariate regression analyses for predictors of COVID-19 death (n=106)

| Characteristic | Death |  |  |  |  |  |  |
| --- | --- | --- | --- | --- | --- | --- | --- |
| | n (%) | $\beta$ | Univariate<br>OR (95% CI) | *P* value | $\beta$ | Multivariate<br>OR (95% CI) | *P* value |
| **Demography** |  |  |  |  |  |  |  |
| Age category, year |  |  |  |  |  |  |  |
| <45 | 0 (0) |  | 1 (ref) |  |  |  |  |
| $\geq 45$ | 7 (100) | 2.81 | 16.6 (1.93 - >999)** | 0.01 | 2.62 | 13.7 (0.87 - >999) | 0.09 |
| Gender |  |  |  |  |  |  |  |
| Female | 3 (42.9) |  | 1 (ref) |  |  |  |  |
| Male | 4 (57.1) | 0.23 | 1.26 (0.27 - 5.9) | 0.77 |  |  |  |
| Travel History |  |  |  |  |  |  |  |
| No | 6 (85.7) |  | 1 (ref) |  |  |  |  |
| Yes | 1 (14.3) | -2.18 | 0.11 (0.01 - 0.98)* | 0.047 | 1.44 | 4.23 (0.25 - 92.9)* | 0.04 |
| History of Contact |  |  |  |  |  |  |  |
| No | 2 (28.6) |  | 1 (ref) |  |  |  |  |
| Yes | 5 (71.4) | 2.72 | 15.2 (2.68 - 86.0) | 0.002 | 4.36 | 78.4 (4.85->999)** | 0.01 |
| **Symptoms** |  |  |  |  |  |  |  |
| Fever |  |  |  |  |  |  |  |
| No | 2 (21.2) |  | 1 (ref) |  |  |  |  |
| Yes | 5 (71.4) | -0.51 | 0.60 (0.13 - 3.54) | 0.53 |  |  |  |
| Cough |  |  |  |  |  |  |  |
| No | 1 (14.3) |  | 1 (ref) |  |  |  |  |
| Yes | 6 (85.7) | 0.15 | 1.16 (0.13 -10.3) | 0.90 |  |  |  |
| Headache |  |  |  |  |  |  |  |
| No | 7 (100) |  | 1 (ref) |  |  |  |  |
| Yes | 0 (0) | -0.33 | 0.72 (0.005 - 6.86) | 0.84 |  |  |  |
| Sore throat |  |  |  |  |  |  |  |
| No | 7 (100) |  | 1 (ref) |  |  |  |  |
| Yes | 0 (0) | -0.67 | 0.51 (0.004 - 4.73) | 0.67 |  |  |  |
| | n (%) | $\beta$ | Univariate<br>OR (95% CI) | *P* value | $\beta$ | Multivariate<br>OR (95% CI) | *P* value |
| Runny Nose |  |  |  |  |  |  |  |
| No | 7 (100) |  | 1 (ref) |  |  |  |  |
| Yes | 0 (0) | -1.37 | 0.25 (0.002-2.25) | 0.37 |  |  |  |
| Dyspnoea |  |  |  |  |  |  |  |
| No | 1 (14.3) |  | 1 (ref) |  |  |  |  |
| Yes | 6 (85.7) | 1.65 | 5.19 (1.04 – 51.1) | 0.08 |  |  |  |
| GI Tract Complains |  |  |  |  |  |  |  |
| No | 7 (100) |  | 1 (ref) |  |  |  |  |
| Yes | 0 (0) | 0.14 | 1.15 (0.008 – 11.9) | 0.94 |  |  |  |
| **Radiology abnormalities** |  |  |  |  |  |  |  |
| Pneumonia |  |  |  |  |  |  |  |
| No | 0 (0) |  | 1 (ref) |  |  |  |  |
| Yes | 7 (100) | 18.9 | 6.58 (0.76-864) | 0.21 |  |  |  |
| Cardiomegaly |  |  |  |  |  |  |  |
| No | 7 (100) |  | 1 (ref) |  |  |  |  |
| Yes | 0 (0) | -0.66 | 0.52 (0.06 - 4.35) | 0.54 |  |  |  |
| **Coexisting diseases** |  |  |  |  |  |  |  |
| Any |  |  |  |  |  |  |  |
| No | 3 (42.9) |  | 1 (ref) |  |  |  |  |
| Yes | 4 (57.1) | 0.64 | 1.89 (0.4 - 8.88) | 0.42 |  |  |  |
| Diabetes Mellitus |  |  |  |  |  |  |  |
| No | 3 (42.9) |  | 1 (ref) |  |  |  |  |
| Yes | 4 (57.1) | 2.47 | 11.9 (2.32 - 60.8)** | 0.003 | 3.57 | 35.6 (2.92->999)* | 0.02 |
| Hypertension |  |  |  |  |  |  |  |
| No | 2 (28.6) |  | 1 (ref) |  |  |  |  |
| Yes | 5 (71.4) | 1.16 | 3.2 (0.55 - 18.5) | 0.19 |  |  |  |
| | n (%) | $\beta$ | Univariate<br>OR (95% CI) | *P* value | $\beta$ | Multivariate<br>OR (95% CI) | *P* value |
| Malignancy |  |  |  |  |  |  |  |
| No | 1 (14.3) |  | 1 (ref) |  |  |  |  |
| Yes | 6 (85.7) | 1.14 | 3.13 (0.31 - 31.3) | 0.33 |  |  |  |
| Heart Disease |  |  |  |  |  |  |  |
| No | 7 (100) |  | 1 (ref) |  |  |  |  |
| Yes | 0 (0) | -0.34 | 0.71 (0.01-6.79) | 0.82 |  |  |  |
| Kidney and Other Diseases |  |  |  |  |  |  |  |
| No | 7 (100) |  | 1 (ref) |  |  |  |  |
| Yes | 0 (0) | -0.46 | 0.63 (0.01-5.91) | 0.77 |  |  |  |
| Respiratory Illness<sup>a</sup> |  |  |  |  |  |  |  |
| No | 7 (100) |  | 1 (ref) |  |  |  |  |
| Yes | 0 (0) | 0.76 | 0.47 (0.004-4.26) | 0.62 |  |  |  |
| Other severe diseases<sup>b</sup> |  |  |  |  |  |  |  |
| No | 1 (14.3) |  | 1 (ref) |  |  |  |  |
| Yes | 6 (85.7) | -1.1 | 0.33 (0.04 - 2.88) | 0.32 |  |  |  |
| **Laboratory Results** |  |  |  |  |  |  |  |
| Anaemia |  |  |  |  |  |  |  |
| No | 3 (50.0) |  | 1 (ref) |  |  |  |  |
| Yes | 3 (50.0) | 0.93 | 2.54 (0.48 - 13.4) | 0.27 |  |  |  |
| Leucocyte |  |  |  |  |  |  |  |
| >10 K | 4 (66.7) |  | 1 (ref) |  |  |  |  |
| <= 10 K | 2 (33.3) | -0.61 | 0.55 (0.10 - 3.13) | 0.50 |  |  |  |
| NLR<sup>c</sup> |  |  |  |  |  |  |  |
| < 3.13 | 0 (0) |  |  |  |  |  |  |
| >= 3.13 | 6 (100) | 0.03 | 1.03 (0.98 – 1.07) | 0.21 |  |  |  |

*NLR* neutrophil-to-lymphocyte ratio.
<sup>a</sup>Included lung TB and other respiratory illness.
<sup>b</sup>Other severe diseases: heart disease, kidney diseases, and severe diseases. <sup>c</sup>based on Liu, et al(7). \*\**p*-values < 0.01; \**p*-values < 0.05

Results for the HCA of symptoms are seen in the dendrogram (Figure 1). HCA cluster 1 (HCA-C1) comprised 7 (38.9%) confirmed, 17 (28.3%) suspected, and 1 (3.6%) lung infection cases. Patients reported predominantly fever with cough, with some also reporting sore throat and GIC, but none with dyspnoea. HCA-C2 comprised 2 (11.1%) confirmed, 11 (18.3%) suspected, and 5 (17.9%) lung infection cases. Patients had predominantly fever, with few combined with dyspnoea, and runny nose. However, cough was not a primary symptom, and none had GIC. Most importantly, patients had fewer symptoms compared to other HCA clusters, and with some reporting no symptoms. HCA-C3 comprised of 17 (28.3%) suspected, but no confirmed or lung infection cases. Patients tended to present with more symptoms, typically 3-5, which were mainly fever, cough, headache, runny nose, and sore throat. A few experienced dyspnoeas and none had GIC. HCA-C4 comprised 1 (5.6%) confirmed, 4 (6.7%) suspected, and 6 (21.4%) lung infection cases. All reported only cough and dyspnoea. HCA-C5 comprised 8 (44.4%) confirmed, 11 (18.3%) suspected and 16 (57%) lung infection cases. All exhibited cough, dyspnoea, and fever.

**Figure 1.**
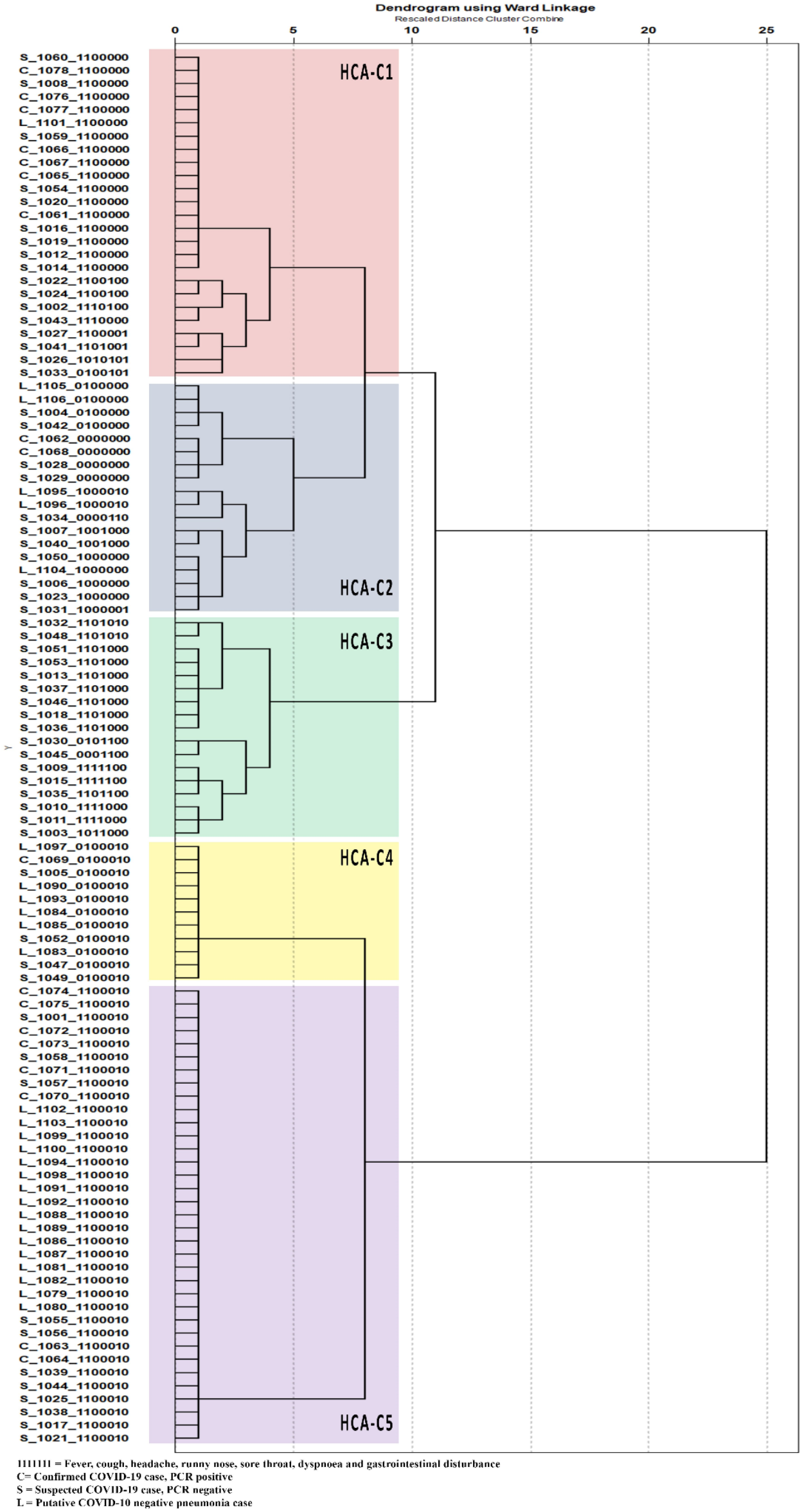
Dendrogram of symptoms cluster for confirmed and suspected COVID-19 cases, and non-COVID-19 pneumonia patients.

The PCA for symptoms revealed two components. PCA symptom 1 (PCA-S1) represented the covariance pattern of patients with headache, runny nose, sore throat, and less dyspnoea with factor loadings of 0.47, 0.43, 0.45, and −0.5, respectively, and accounted for 30% of the total variance across symptoms. PCA-S2 represented a pattern of fever, cough, and GIC with factor loadings of 0.65, 0.51 and 0.63, respectively, and accounted for 18% of variance. The PCA for haematology revealed three components. PCA-H1 represented haemoglobin and haematocrit with loadings of 0.54 and 0.52 and accounted for 42% of total variance. PCA-H2 represented lymphocyte and neutrophil counts with loadings of 0.76 and − 0.83 and accounted for 18% of variance. PCA-H3 had a high factor loading of 1.15 for monocytes and tended toward lower factor loadings of −0.05, −0.05, −0.02, 0.09, −0.12, −0.05, 0.13 and −0.05 for haemoglobin, haematocrit, thrombocytes, leucocytes, lymphocytes, basophils, neutrophils, and eosinophils, respectively, and accounted for 14% of total variance.

Univariate logistic regression of symptom data on SARS-CoV-2 infection revealed that runny nose tended toward decreased risk (OR 0.1; 95%CI < 0.001 – 0.83; p = 0.08). No other symptoms exhibited associations in either univariate or multivariate analysis (Table 4). Multivariate analysis of HCA groups revealed that HCA-C3 tended toward reduced risk of COVID-19 cases (aOR 0.048; 95%CI < 0.01 – 0.52; p = 0.056). HCA-C3 comprised patients with symptoms of fever, cough, headache, runny nose, or sore throat, in which only a few experienced dyspnoea, and none had GIC. No other clusters showed any significant associations with COVID-19 cases. However, in all multivariate models with HCA predictors, age ≥ 45 years, presence of international travel history, previous contact with a COVID-19 patient, and pneumonia by x-ray were significant associations, consistent with previous results. For mortality, none of the symptoms nor HCA clusters showed any association in univariate or multivariate analyses (Table 5). However, the presence of diabetes and history of contact with COVID-19 patients were consistently associated with higher mortality in all HCA models.

**Table 4.**
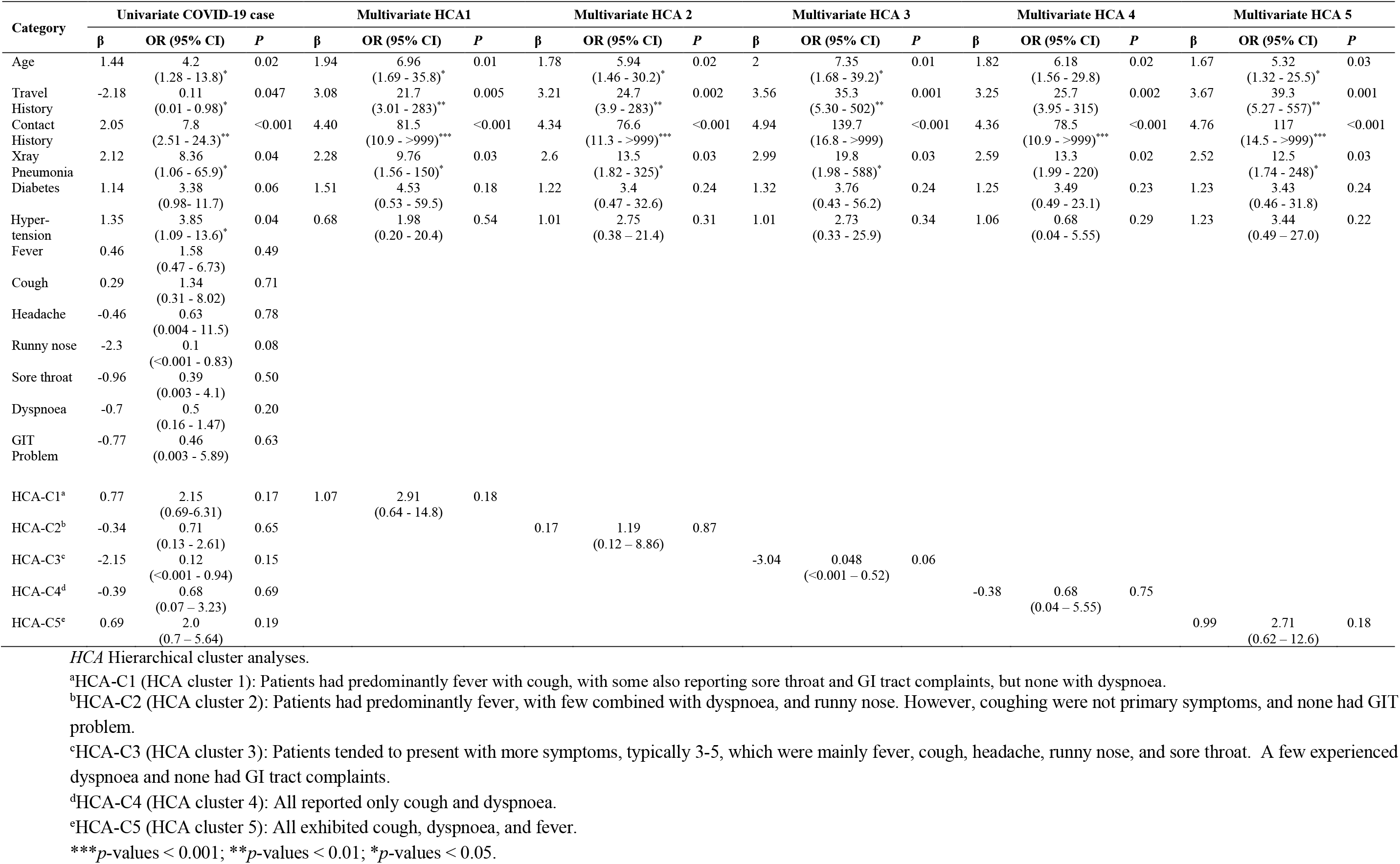
Univariate and multivariate logistic regression analyses of HCA clusters and SARS-CoV-2 infection (n=106)

**Table 5.** Univariate and multivariate logistic regression analyses of HCA clusters and COVID-19 death (n=106)

| Category | Univariate COVID-19 Death |  |  | Multivariate (HCA1) |  |  | Multivariate HCA 2 |  |  | Multivariate HCA 3 |  |  | Multivariate HCA 4 |  |  | Multivariate HCA 5 |  |  |
| --- | --- | --- | --- | --- | --- | --- | --- | --- | --- | --- | --- | --- | --- | --- | --- | --- | --- | --- |
| | $\beta$ | OR (95% CI) | P | $\beta$ | OR (95% CI) | P | $\beta$ | OR (95% CI) | P | $\beta$ | OR (95% CI) | P | $\beta$ | OR (95% CI) | P | $\beta$ | OR (95% CI) | P |
| Age | 2.81 | 16.6<br>(1.93 - >999) | 0.01 | 2.74 | 15.4<br>(0.67 - >999) | 0.12 | 1.98 | 7.23<br>(0.54 - >999) | 0.17 | 2.0 | 7.41<br>(0.49 - >999) | 0.17 | 1.69 | 5.44<br>(0.41 - 767) | 0.22 | 1.96 | 7.11<br>(0.51 - >999) | 0.18 |
| Travel History | -2.18 | 0.11<br>(0.01 - 0.98)* | 0.047 | 2.44 | 11.5<br>(0.50 - >999) | 0.12 | 1.41 | 4.09<br>(0.27 - 78.1) | 0.28 | 1.43 | 4.18<br>(0.28 - 75.3) | 0.27 | 1.03 | 2.8<br>(0.16 - 47.2) | 0.42 | 2.23 | 9.27<br>(0.42 - >999) | 0.13 |
| Contact History | 2.72 | 15.2<br>(2.68 - 86.0) | 0.002 | 4.97 | 144<br>(6.45 - >999)* | 0.004 | 4.01 | 55.0<br>(4.32 - >999)* | 0.005 | 4.26 | 70.5<br>(5.14 - >999)** | 0.004 | 3.93 | 51<br>(4.28 - >999)** | 0.004 | 4.78 | 118.7<br>(5.16 - >999)** | 0.01 |
| X-ray Pneumonia | 18.9 | 6.58<br>(0.76 - 864) | 0.21 | 1.79 | 6.01<br>(0.35 - >999) | 0.26 | 1.69 | 5.44<br>(0.33 - >999)* | 0.29 | 1.28 | 3.6<br>(0.22 - 651) | 0.39 | 1.32 | 3.73<br>(0.26 - 546) | 0.37 | 1.14 | 3.12<br>(0.20 - 545) | 0.44 |
| Diabetes | 2.47 | 11.9<br>(2.32 - 60.8)** | 0.003 | 3.09 | 22.0<br>(1.44 - >999)* | 0.039 | 3.19 | 24.4<br>(2.1 - >999) | 0.02 | 3.15 | 23.4<br>(1.83 - >999)* | 0.03 | 2.95 | 19<br>(1.75 - >999)* | 0.03 | 3.45 | 31.4<br>(1.84 - >999)* | 0.02 |
| Hypertension | 1.16 | 3.2<br>(0.55 - 18.5) | 0.19 | 1.13 | 3.09<br>(0.12 - 490) | 0.45 | -0.16 | 0.85<br>(0.055 - 8.31) | 0.89 | -0.41 | 0.67<br>(0.04 - 6.37) | 0.74 | -0.19 | 0.83<br>(0.07 - 7.76) | 0.87 | -0.05 | 3.88<br>(0.45 - 66.5) | 0.97 |
| Fever | -0.37 | 0.69<br>(0.15 - 4.14) | 0.64 |  |  |  |  |  |  |  |  |  |  |  |  |  |  |  |
| Cough | -0.71 | 0.49<br>(0.061 - 5.16) | 0.45 |  |  |  |  |  |  |  |  |  |  |  |  |  |  |  |
| Headache | 0.84 | 2.32<br>(0.012 - 125) | 0.61 |  |  |  |  |  |  |  |  |  |  |  |  |  |  |  |
| Runny nose | -0.41 | 0.67<br>(0.005 - 7.46) | 0.75 |  |  |  |  |  |  |  |  |  |  |  |  |  |  |  |
| Sore throat | -0.34 | 0.71<br>(0.002 - 13.1) | 0.82 |  |  |  |  |  |  |  |  |  |  |  |  |  |  |  |
| Dyspnoea | 1.18 | 3.26<br>(0.61 - 35.5) | 0.18 |  |  |  |  |  |  |  |  |  |  |  |  |  |  |  |
| GIT Problem | 0.69 | 2.0<br>(0.007 - 47.6) | 0.68 |  |  |  |  |  |  |  |  |  |  |  |  |  |  |  |
| HCA-C1<sup>a</sup> | -1.6 | 0.2<br>(0.002 - 1.77) | 0.29 | 2.84 | 0.06<br>(<0.001 - 2.02) | 0.17 |  |  |  |  |  |  |  |  |  |  |  |  |
| HCA-C2<sup>b</sup> | 0.072 | 1.07<br>(0.11 - 5.6) | 0.94 |  |  |  | 0.87 | 2.4<br>(0.16 - 45.5) | 0.48 |  |  |  |  |  |  |  |  |  |
| HCA-C3<sup>c</sup> | -1.17 | 0.31<br>(0.002 - 2.76) | 0.44 |  |  |  |  |  |  | -1.35 | 0.26<br>(0.002 - 3.83) | 0.4 |  |  |  |  |  |  |
| HCA-C4<sup>d</sup> | 0.67 | 1.95<br>(0.19 - 10.7) | 0.51 |  |  |  |  |  |  |  |  |  | 1.07 | 2.93<br>(0.13 - 51.5) | 0.44 |  |  |  |
| HCA-C5<sup>e</sup> | 1.6 | 4.94<br>(1.12 - 28.8)* | 0.046 |  |  |  |  |  |  |  |  |  |  |  |  | 1.36 | 3.88<br>(0.45 - 66.5) | 0.20 |

*HCA* Hierarchical cluster analyses.
<sup>a</sup>HCA-C1 (HCA cluster 1): Patients had predominantly fever with cough, with some also reporting sore throat and GI tract complaints, but none with dyspnoea.
<sup>b</sup>HCA-C2 (HCA cluster 2): Patients had predominantly fever, with few combined with dyspnoea, and runny nose. However, coughing were not primary symptoms, and none had GIT problem.
<sup>c</sup>HCA-C3 (HCA cluster 3): Patients tended to present with more symptoms, typically 3-5, which were mainly fever, cough, headache, runny nose, and sore throat. A few experienced dyspnoea and none had GI tract complaints.
<sup>d</sup>HCA-C4 (HCA cluster 4): All reported only cough and dyspnoea.
<sup>e</sup>HCA-C5 (HCA cluster 5): All exhibited cough, dyspnoea, and fever.
**\*\****p*-values < 0.01; **\****p*-values < 0.05.

Analysis of PCA revealed a symptom component for fever, cough, and GIC that tended to increase risk of infection (OR 3.41; 95% CI 1.06-14; p=0.06), and a haematology component with elevated monocytes decreased risk (OR 0.26; 95%CI 0.07-0.79; p=0.027) (Table 6). Multivariate models that included either PCA clusters or the individual predictors were consistent in that presence of travel history and contact with COVID-19 patients were significant predictors of COVID-19 cases.

**Table 6.** Univariate and multivariate logistic regression analyses of PCA components and SARS-CoV-2 infection (n=106)

| Category | Univariate COVID-19 case |  |  | Multivariate PCA Symptom & Haematology |  |  | Multivariate PCA Symptom |  |  | Multivariate PCA Haematology |  |  |
| --- | --- | --- | --- | --- | --- | --- | --- | --- | --- | --- | --- | --- |
| | $\beta$ | OR (95% CI) | P | $\beta$ | aOR (95% CI) | P | $\beta$ | aOR (95% CI) | P | $\beta$ | aOR (95% CI) | P |
| Age | 1.44 | 4.20 (1.28 - 13.8)* | 0.02 | 2.03 | 7.58 (1.36 - 61.6) | 0.03 | 2.20 | 9.01 (1.73 - 65.8) | 0.01 | 1.66 | 5.26 (1.24 - 27.8)* | 0.03 |
| Travel History | -2.18 | 0.11 (0.01 - 0.98)* | 0.047 | 2.73 | 15.4 (1.65 - >999) | 0.02 | 2.97 | 19.5 (3.21 - 216) | 0.003 | 3.07 | 21.5 (2.60 - 387)** | 0.01 |
| Contact History | 2.05 | 7.80 (2.51 - 24.3)*** | <0.001 | 4.00 | 54.8 (5.08 - >999) | 0.002 | 4.21 | 67.5 (9.13 - >999) | <0.001 | 4.07 | 58.6 (7.49 - >999)*** | <0.001 |
| Xray Pneumonia | 2.12 | 8.36 (1.06 - 65.9)* | 0.04 | 1.28 | 3.60 (0.38 - 84.8) | 0.24 | 2.06 | 7.87 (0.89 - 168) | 0.08 | 1.70 | 5.46 (0.99 - 59.1) | 0.07 |
| Diabetes | 1.14 | 3.38 (0.98 - 11.7) | 0.06 | 0.77 | 2.13 (0.16 - 72.7) | 0.51 | 1.04 | 2.82 (0.28 - 39.4) | 0.36 | 1.16 | 3.19 (0.24 - 69.6) | 0.34 |
| Hypertension | 1.35 | 3.85 (1.09 - 13.6)* | 0.04 | 0.69 | 1.99 (0.05 - 82.9) | 0.62 | 0.22 | 1.25 (0.09 - 14.8) | 0.85 | 0.99 | 2.68 (0.19 - 47.0) | 0.41 |
| Fever | 0.46 | 1.58 (0.47 - 6.73) | 0.49 |  |  |  |  |  |  |  |  |  |
| Cough | 0.29 | 1.34 (0.31 - 8.02) | 0.71 |  |  |  |  |  |  |  |  |  |
| Headache | -0.46 | 0.63 (0.004 - 11.5) | 0.78 |  |  |  |  |  |  |  |  |  |
| Runny nose | -2.30 | 0.10 (<0.001 - 0.83) | 0.08 |  |  |  |  |  |  |  |  |  |
| Sore throat | -0.96 | 0.39 (0.003 - 4.10) | 0.50 |  |  |  |  |  |  |  |  |  |
| Dyspnoea | -0.70 | 0.50 (0.16 - 1.47) | 0.20 |  |  |  |  |  |  |  |  |  |
| GIT Problem | -0.77 | 0.46 (0.003 - 5.89) | 0.63 |  |  |  |  |  |  |  |  |  |
| PCA-S1 (1)<sup>a</sup> | 0.58 | 1.79 (0.57 - 5.59) | 0.32 | 0.28 | 1.32 (0.11 - 12.8) | 0.79 | 0.84 | 2.31 (0.36 - 19.3) | 0.37 |  |  |  |
| PCA-S1 (2)<sup>b</sup> | -1.10 | 0.33 (0.06 - 1.30) | 0.15 | 0.51 | 1.67 (0.05 - 63.6) | 0.72 | 0.21 | 1.23 (0.06 - 23.4) | 0.88 |  |  |  |
| PCA-S2 (1)<sup>c</sup> | 1.23 | 3.41 (1.06 - 14.0) | 0.06 | 1.13 | 3.10 (0.24 - 177) | 0.31 | 0.69 | 1.99 (0.27 - 25.7) | 0.49 |  |  |  |
| PCA-S2 (2)<sup>d</sup> | -1.33 | 0.27 (0.002 - 2.97) | 0.40 | -2.74 | 0.06 (<0.001 - 1.70) | 0.13 | -2.62 | 0.07 (<0.001 - 1.48) | 0.14 |  |  |  |
| PCA-H1 (1)<sup>e</sup> | 0.06 | 1.07 (0.25 - 4.48) | 0.93 | -0.09 | 0.91 (0.10 - 9.64) | 0.92 |  |  |  | 0.13 | 1.14 (0.18 - 8.10) | 0.88 |
| PCA-H1 (2)<sup>f</sup> | 1.05 | 2.86 (0.89 - 10.6) | 0.10 | 0.94 | 2.56 (0.34 - 29.8) | 0.32 |  |  |  | 0.75 | 2.13 (0.43 - 12.3) | 0.35 |
| PCA-H2 (1)<sup>g</sup> | -0.96 | 0.39 (0.09 - 1.39) | 0.17 | -1.24 | 0.29 (0.02 - 2.81) | 0.24 |  |  |  | -1.39 | 0.25 (0.03 - 1.69) | 0.15 |
| PCA-H2 (2)<sup>h</sup> | -0.19 | 0.82 (0.27 - 2.51) | 0.74 | 0.42 | 1.53 (0.20 - 3.04) | 0.66 |  |  |  | -0.54 | 0.58 (0.09 - 3.44) | 0.53 |
| PCA-H3 (1)<sup>i</sup> | -1.36 | 0.26 (0.07 - 0.79)* | 0.03 | -0.34 | 0.71 (0.11 - 4.75) | 0.69 |  |  |  | -0.79 | 0.45 (0.08 - 2.19) | 0.33 |
| PCA-H3 (2)<sup>j</sup> | -1.83 | 0.16 (0.03 - 0.61)* | 0.02 | -1.29 | 0.28 (0.01 - 3.47) | 0.26 |  |  |  | -1.13 | 0.32 (0.03 - 2.05) | 0.23 |

PCA Principal component analyses.
<sup>a</sup>PCA-S1 (1) (PCA Symptom 1, 2<sup>nd</sup> tertile vs 1<sup>st</sup> tertile): Headache, runny nose, sore throat, and less dyspnoea with factor loadings of 0.47, 0.43, 0.45, and -0.5, respectively.
<sup>b</sup>PCA-S1 (2) (PCA Symptom 1, 3<sup>rd</sup> tertile vs 1<sup>st</sup> tertile): Headache, runny nose, sore throat, and less dyspnoea with factor loadings of 0.47, 0.43, 0.45, and -0.5, respectively.
<sup>c</sup>PCA-S2 (1) (PCA Symptom 2, 2<sup>nd</sup> tertile vs 1<sup>st</sup> tertile): Fever, cough, and GI tract complaints with factor loadings of 0.65, 0.51 and 0.63, respectively.
<sup>d</sup>PCA-S2 (2) (PCA Symptom 2, 3<sup>rd</sup> tertile vs 1<sup>st</sup> tertile): Fever, cough, and GI tract complaints with factor loadings of 0.65, 0.51 and 0.63, respectively.
<sup>e</sup>PCA-H1 (1) (PCA Haematology 1, 2<sup>nd</sup> tertile vs 1<sup>st</sup> tertile): Haemoglobin and haematocrit with factor loadings of 0.54 and 0.52, respectively.
<sup>f</sup>PCA-H1 (2) (PCA Haematology 1, 3<sup>rd</sup> tertile vs 1<sup>st</sup> tertile): Haemoglobin and haematocrit with factor loadings of 0.54 and 0.52, respectively.
<sup>g</sup>PCA-H2 (1) (PCA Haematology 2, 2<sup>nd</sup> tertile vs 1<sup>st</sup> tertile): Lymphocyte and neutrophil counts with factor loadings of 0.76 and -0.83, respectively.
<sup>h</sup>PCA-H2 (2) (PCA Haematology 2, 3<sup>rd</sup> tertile vs 1<sup>st</sup> tertile): Lymphocyte and neutrophil counts with factor loadings of 0.76 and -0.83, respectively.

In univariate analyses for death, HCA-C5 of cough, fever and dyspnoea had increased risk (OR 5.75; P5%CI 1.06 − 31.3, p=0.043). No other individual symptoms or haematology measures, nor HCA clusters or PCA components were associated with death in univariate or multivariate analyses (Table 7). However, we note that diabetes tended toward increased risk for death (aOR 8.86; 95%CI 0.72 – 17.0; p = 0.07), and that history of contact with COVID-19 cases was consistently associated with death (aOR 28.6; 95%CI 1.63->999.9; p = 0.01).

**Table 7.** Univariate and multivariate logistic regression analyses of PCA components and COVID-19 death (n=106)

| Category | Univariate COVID-19 death |  |  | Multivariate PCA Symptom & Haematology |  |  | Multivariate PCA Symptom |  |  | Multivariate PCA Haematology |  |  |
| --- | --- | --- | --- | --- | --- | --- | --- | --- | --- | --- | --- | --- |
| | $\beta$ | OR (95% CI) | P | $\beta$ | OR*(95% CI) | P | $\beta$ | OR*(95% CI) | P | $\beta$ | OR*(95% CI) | P |
| Age | 2.81 | 16.6 (1.93 - >999)** | 0.01 | 1.77 | 5.86 (0.40 - > 999) | 0.14 | 1.84 | 6.28 (0.21 - >999) | 0.23 | 2.85 | 17.4 (0.68 - >999) | 0.08 |
| Travel History | -2.18 | 0.11 (0.01 - 0.98)* | 0.047 | 1.15 | 3.15 (0.04 - 613) | 0.38 | 1.81 | 6.08 (0.31 - >999) | 0.17 | 1.60 | 4.97 (0.12 - >999) | 0.28 |
| Contact History | 2.72 | 15.2 (2.68 - 86.0)** | 0.002 | 3.35 | 28.6 (1.63 - > 999)** | 0.01 | 4.43 | 84.2 (5.66 - >999)** | 0.002 | 3.81 | 44.9 (2.35 - 87.6)** | 0.01 |
| Xray Pneumonia | 18.9 | 6.58 (0.76 - 864) | 0.21 | 1.05 | 2.86 (0.08 - 377) | 0.41 | 2.09 | 8.08 (0.27 - >999) | 0.21 | 0.96 | 2.62 (0.002 - >999) | 0.51 |
| Diabetes | 2.47 | 11.9 (2.32 - 60.8)** | 0.003 | 2.18 | 8.86 (0.72 - 17.0) | 0.07 | 2.30 | 10.0 (0.82 - >999) | 0.07 | 3.04 | 20.8 (1.27 - >999)* | 0.03 |
| Hypertension | 1.16 | 3.2 (0.55 - 18.5) | 0.19 | 0.36 | 1.43 (0.08 - 64.8) | 0.76 | 0.67 | 1.95 (0.12 - 192) | 0.60 | 0.35 | 1.42 (0.047 - 582) | 0.79 |
| Fever | -0.37 | 0.69 (0.15 - 4.14) | 0.64 |  |  |  |  |  |  |  |  |  |
| Cough | -0.71 | 0.49 (0.06 - 5.16) | 0.45 |  |  |  |  |  |  |  |  |  |
| Headache | 0.84 | 2.32 (0.01 - 125) | 0.61 |  |  |  |  |  |  |  |  |  |
| Runny nose | -0.41 | 0.67 (0.005 - 7.46) | 0.75 |  |  |  |  |  |  |  |  |  |
| Sore throat | -0.34 | 0.71 (0.002 - 13.1) | 0.82 |  |  |  |  |  |  |  |  |  |
| Dyspnoea | 1.18 | 3.26 (0.61 - 35.5) | 0.18 |  |  |  |  |  |  |  |  |  |
| GIT Problem | 0.69 | 2.0 (0.007 - 47.6) | 0.68 |  |  |  |  |  |  |  |  |  |
| PCA-S1 (1)<sup>a</sup> | -2.13 | 0.12 (<0.001 - 1.08) | 0.16 | -1.70 | 0.18 (< 0.001 - 0.36) | 0.25 | -2.37 | 0.09 (<0.001 - 1.76) | 0.14 |  |  |  |
| PCA-S1 (2)<sup>b</sup> | -1.33 | 0.26 (0.03 - 1.35) | 0.16 | -0.07 | 0.51 (< 0.001 - 65.8) | 0.66 | -0.70 | 0.49 (0.01 - 108) | 0.67 |  |  |  |
| PCA-S2 (1)<sup>c</sup> | 0.42 | 1.52 (0.34 - 8.86) | 0.60 | -0.35 | 0.71 (0.03 - 21.8) | 0.76 | -0.48 | 0.62 (0.04 - 38.4) | 0.70 |  |  |  |
| PCA-S2 (2)<sup>d</sup> | -0.96 | 0.38 (0.003 - 5.05) | 0.55 | 0.85 | 2.33 (< 0.001 - 204) | 0.56 | -1.02 | 0.36 (<0.001 - 15.8) | 0.59 |  |  |  |
| PCA-H1 (1)<sup>e</sup> | -0.31 | 0.74 (0.12 - 4.04) | 0.73 | 1.03 | 2.79 (0.15 - 5.43) | 0.33 |  |  |  | 1.11 | 3.04 (0.16 - >999) | 0.36 |
| PCA-H1 (2)<sup>f</sup> | -0.37 | 0.69 (0.11 - 3.79) | 0.68 | -0.43 | 0.65 (0.002 - 19.8) | 0.73 |  |  |  | -1.58 | 0.21 (<0.001 - 5.18) | 0.29 |
| PCA-H2 (1)<sup>g</sup> | -0.79 | 0.46 (0.08 - 2.05) | 0.33 | -0.07 | 0.93 (0.06 - 18.3) | 0.94 |  |  |  | -0.07 | 0.93 (0.04 - 36.9) | 0.96 |
| PCA-H2 (2)<sup>h</sup> | -2.57 | 0.08 (<0.001 - 0.72) | 0.09 | -1.30 | 0.27 (<0.001 - 0.54) | 0.26 |  |  |  | -1.81 | 0.16 (<0.001 - 2.42) | 0.18 |
| PCA-H3 (1)<sup>i</sup> | -0.94 | 0.39 (0.07 - 1.76) | 0.25 | -1.53 | 0.22 (<0.001 - 3.09) | 0.22 |  |  |  | -1.78 | 0.17 (<0.001 - 3.84) | 0.20 |
| PCA-H3 (2)<sup>j</sup> | -2.43 | 0.09 (<0.001 - 0.83) | 0.11 | -1.07 | 0.34 (0.005 - 5.33) | 0.35 |  |  |  | -1.82 | 0.16 (<0.001 - 2.15) | 0.17 |

PCA Principal component analyses.
<sup>a</sup>PCA-S1 (1) (PCA Symptom 1, 2<sup>nd</sup> tertile vs 1<sup>st</sup> tertile): Headache, runny nose, sore throat, and less dyspnoea with factor loadings of 0.47, 0.43, 0.45, and -0.5, respectively.
<sup>b</sup>PCA-S1 (2) (PCA Symptom 1, 3<sup>rd</sup> tertile vs 1<sup>st</sup> tertile): Headache, runny nose, sore throat, and less dyspnoea with factor loadings of 0.47, 0.43, 0.45, and -0.5, respectively.
<sup>c</sup>PCA-S2 (1) (PCA Symptom 2, 2<sup>nd</sup> tertile vs 1<sup>st</sup> tertile): Fever, cough, and GI tract complaints with factor loadings of 0.65, 0.51 and 0.63, respectively.
<sup>d</sup>PCA-S2 (2) (PCA Symptom 2, 3<sup>rd</sup> tertile vs 1<sup>st</sup> tertile): Fever, cough, and GI tract complaints with factor loadings of 0.65, 0.51 and 0.63, respectively.
<sup>e</sup>PCA-H1 (1) (PCA Haematology 1, 2<sup>nd</sup> tertile vs 1<sup>st</sup> tertile): Haemoglobin and haematocrit with factor loadings of 0.54 and 0.52, respectively.
<sup>f</sup>PCA-H1 (2) (PCA Haematology 1, 3<sup>rd</sup> tertile vs 1<sup>st</sup> tertile): Haemoglobin and haematocrit with factor loadings of 0.54 and 0.52, respectively.
<sup>g</sup>PCA-H2 (1) (PCA Haematology 2, 2<sup>nd</sup> tertile vs 1<sup>st</sup> tertile): Lymphocyte and neutrophil counts with factor loadings of 0.76 and -0.83, respectively.
<sup>h</sup>PCA-H2 (2) (PCA Haematology 2, 3<sup>rd</sup> tertile vs 1<sup>st</sup> tertile): Lymphocyte and neutrophil counts with factor loadings of 0.76 and -0.83, respectively.

## Discussion

To our knowledge, this is the first report of confirmed and suspected cases of COVID-19 in Indonesia. It is unique in examining patterns, clusters, or covariance constructs of symptoms and haematological data, or combinations thereof, with other predictors and their associations with key outcomes of infection and death.

The present study found consistent results in multivariate analyses for age ≥ 45 years, presence of travel history, contact with a COVID-19 patient, and pneumonia by x-ray as significant predictors of COVID-19 cases. The associations persisted when adjusted with data for symptoms and haematology either individually or as clusters or components. In contrast, analysis of individual symptoms in either univariate or multivariate analysis did not reveal any associations, but HCA yielded a cluster that tended toward reduced risk of COVID-19 cases, and included patients that typically exhibited 3-5 symptoms, which were mainly fever, cough, headache, runny nose, or sore throat, few GIC, and lacking dyspnoea. Other PCA components tended toward associations in univariate analysis, although none was significant, nor were any significant in multivariate analysis. Nevertheless, the stronger tendency for associations of cluster and covariance constructs underscore the importance of interpreting infection as a systemic event.

Our results are in line with previous studies that identified older age, history of exposure, chronic underlying disease, symptoms, laboratory findings and chest x-ray examination. Specifically, we observed that persons ≥ 45 years had increased risk of infection, with 75% of confirmed cases being 45-65 years of age. This echoes previous reports wherein mean age of patients was over 40 years, and where critically ill patients were older than non-critical patients.(7) Liu W, et al., showed that age was significantly associated with the COVID-19 cases (OR, 10.6; 95% CI: 2.10– 53.4; p value = 0.004) and disease progression (OR, 8.55; 95% CI: 1.63– 44.8; P = 0.011) (8), similar to our study with aORs ranging from 5.32 – 9.01.

We also showed travel history increased the risk of COVID-19 cases with aORs 2.8 − 19.5, and with travel to Singapore or Malaysia being the most common endemic countries recently visited. This is supported by findings from Wells CR, et al. wherein the daily risk of exporting a confirmed case via international travel exceeded 95% by January 13, 2020 (9). They estimated that 64.3% of exported cases were pre-symptomatic while travelling. As expected, we observed that reported history of contact with a COVID-19 case was a very strong predictor of infection (OR 232; 95% CI 15.1-3569), similar to Liang W-hua, et al., where 1,334 of 1,590 (83.9%) had exposure history (10), and Wang R, et al. where 57.6% of patients had a history of exposure to persons from Wuhan within the past 2 weeks.(11) However, the association of recall of exposure with increased mortality was unanticipated, and warrants further exploration.

Our observation that pneumonia by chest x-ray strongly associated with infection with aORs 3.15 – 19.8 would be expected, and seen in previous report from Fuyang, China where 80% patients showed bilateral pneumonia by chest x-ray, and all of the critical patients showed pneumonia by x-ray.(11)

Similar to other findings, fever, cough and dyspnoea occurred in most patients, with cough and fever associated with infection, although not significant. Univariate regression with cluster analyses confirmed that patients with multiple symptoms, but without dyspnoea, were less likely to be infected (OR 0.04; 95% CI: <0.00 –0.93; P= 0.037). This is consistent with reports that respiratory failure is associated with infection (OR, 8.021; 95% CI: 2.022– 31.821; P = 0.003).(8)

The results on GIC warrant further study as angiotensin converting enzyme 2 (ACE-2), a virus receptor, is expressed in gastrointestinal epithelial cells. Viral nucleocapsid protein has also been identified in the gastric, duodenal, and rectum glandular epithelial cell cytoplasm. GI infection by SARS-CoV-2 may be important, and GI symptoms should not be overlooked among suspected cases.

We did not observe any significant associations of any laboratory results with infection or death. We note that a high neutrophil-to-lymphocyte ratio (NLR) with cut-off ≥ 3.13 was associated with a non-significant increased risk of infection. This cut-off value was based on previous studies which highlighted the importance of the NLR which reflects systemic inflammation.(7) Other than mortality, we did not explore the association of markers on with clinical severity of illness as evaluated in previous studies.

We found diabetes was a consistent and strong predictor for COVID-19 death. Diabetes was previously reported as a risk factor for severe disease and mortality in SARS and MERS (Middle East respiratory syndrome) coronavirus infections, and H1N1 influenza. A recent review summarized that diabetes increased the risk for SARS-CoV-2 infection and hospitalization, with a two-fold risk of severe disease or admission to the intensive care unit (ICU), and increased risk of death.(12) In our study 28% of COVID-19 patients had diabetes or hypertension. Others report increased comorbidity of cardiovascular, endocrine, and digestive disease, of which hypertension (30%) and diabetes (12.1%) were the most common.(13-16)

For death, we observed age ≥ 45 years tended toward an association but was not consistent across analyses. A study by Zhou, et al., showed that age is a predictor for mortality. (17) We also report no significant difference in the risk for infection or death based on sex. As mentioned, reported contact history was a predictor for mortality with aORs 51 – 144. One study of 3,019 suspected cases among health workers documented 1,716 confirmed cases with nearly 15% being severe and with 5 deaths. It was noted that transmission occurred mostly among close contacts.(18) The underlying mechanism for how diabetes and older age are associated with adverse outcomes remains unclear, a current study of case series of COVID-19 patients demonstrated the potential contribution of pre-existing endothelial dysfunction. Post-mortem analysis of a renal transplant recipient aged 71 years whose condition deteriorated after COVID-19 diagnosis showed structures of viral inclusion in endothelial cells and accumulation of endothelium-associated inflammatory cells. And post-mortem analysis of a 58-year diabetic woman revealed lymphocytic endotheliosis in lung, heart, kidney, and liver, in addition to evidence of myocardial infarction, but without signs of lymphocytic myocarditis. Endothelial cells express the ACE-2 virus receptor(19), and there may be increased expression of ACE-2 in the presence of comorbidities.(20-22) Hence, COVID-19 could cause widespread endothelial dysfunction from direct infection or immune-mediated recruitment of inflammatory cells, and provoke multiple organ failure. However, the relatively small size of our study precluded meaningful analysis of comorbidities.

Indonesia hosts many popular tourist destinations which created logistic, economic and political challenges to limiting foreign visitors. As part of the outbreak response, the Ministry of Health ordered the use of thermal scanners at ports of entry on 5 February 2020, prepared approximately 100 hospitals as COVID-19 referral hospitals, and assigned the National Institute for Health Research and Development to establish a coronavirus diagnostic laboratory supported by WHO and the USA Centers for Disease Control. The first two positive cases in Indonesia were announced by the President on 2 March 2020, but the actual number of cases remains unknown due to lack of active surveillance, lags in testing, and a testing strategy that limits assessment of patients with mild symptoms or persons asymptomatic for respiratory infection.

Indonesia now focuses on monitoring and preventing further spread by isolation, self-quarantine, and social distancing especially in large cities, such as Jakarta and Surabaya.(1, 23) The predictors in our study for increased risk of COVID-19 cases (age, international travel history, contact with a COVID-19 patient, pneumonia diagnosis by x-ray) and higher mortality (presence of diabetes and contact history), can be used as essential data for screening and surveillance, and help guide action to reduce transmission and mitigate effects from a possible second wave of COVID-19.

There are limitations to our study. First, the sample size was small as this is a preliminary study on COVID-19 during its emergence in Indonesia, and was limited to data that were accessible from that period. We also note the lack of data on smoking and body mass index. This is may be important because ACE-2 expression has been reported to increase in smokers and persons with complications of high body mass index.(24-26) Further investigation of correlates of expression, activation, and interaction of ACE-2 with SARS-CoV-2 in the Indonesian population would be important.

The likelihood of underreported cases needs to be considered given the variety of clinical manifestations and suboptimal availability of tools for diagnosis. Actions need to be taken such as a simulation to assess and verify pandemic preparedness for influx of cases at referral hospitals for proper triage, and establishment of high throughput diagnostics laboratories for COVID-19.

## Conclusions

In independent and clustered variables analysis, older age, international travel history, contact with a COVID-19 patient, and pneumonia by x-ray were the predictors for increased risk of SARS-CoV-2 infection. The presence of diabetes and history of contact with COVID-19 cases were consistently associated with higher mortality. Association of individual symptoms with infection or mortality were not observed, whereas clustered symptoms or components tended to show stronger associations with infection. Use of such clusters in patient screening and assessment will be useful at the clinic and community level. The Ministry of Health needs to focus on preparedness measures, surveillance and public information to prevent movement of SARS-CoV-2 infection, and create messaging to alleviate public anxiety to enhance action and responsible behaviours.

## Data Availability

The datasets used and/or analysed during the current study are available from the corresponding author on reasonable request.

## List of abbreviations

ACE-2: angiotensin converting enzyme 2
COVID-19: Coronavirus disease 2019
CT: computed tomography
GIC: gastrointestinal complaints
HCA: hierarchical cluster analyses
IQR: interquartile range
NIHRD: National Institutes for Health Research and Development
NLR: neutrophil-to-lymphocyte ratio
PCA: principal component analyses
RT-PCR: real time polymerase chain reaction
SARS-CoV-2: Severe acute respiratory syndrome coronavirus 2
SD: standard deviation
TB: lung tuberculosis

## Declarations

### Ethical approval and consent to participate

Ethical approval was obtained from the Research Ethics Committee of the Faculty of Medicine, Universitas Indonesia – Dr. Cipto Mangunkusumo General Hospital, with approval number 20030331. After consideration of ethical issues, logistics and urgency of this work, the Committee waived the requirement for written individual informed consent and approved the sharing of anonymized data.

## Consent for publication

Not applicable.

## Authors’ contributions

RA, AJR, AFS, DO, EB and AHS designed and coordinated this study. EB, RA, AG, KQP, IGS, P, FI, DP, DL, NGL, VS and ADS collected the data. The data was analysed by RA and interpreted by EB, RA, ISW, and AHS. EB, RA, AHS, ISW, AG, KQP and DR drafted the manuscript together with the help of AJR. AHS provided feedback and helped streamline the content and design analytical procedures. The corresponding author RA coordinated the final submission of the paper. All authors contributed to the final version of the manuscript.

## Acknowledgements

We thank Diah Handayani, Andriansjah Rukmana, Muhammad Luqman Labib Zufar, Prasetio Adinugroho, and Annisa Feby Canintika for their insights and outstanding support in drafting this study. We acknowledge the contributions from COVID-19 team of Persahabatan Hospital, Sanglah Hospital, Gatot Subroto Central Army Hospital, and Raden Mattaher Hospital to several authors and contributors to this work. This study was supported by the in kind contributions of the Dean’s Office and the Indonesian Medical Education and Research Institute of the Faculty of Medicine Universitas Indonesia.

